# Using electronic health records to evaluate a children and young people’s social prescribing service: Challenges and implications for research and practice

**DOI:** 10.64898/2025.12.17.25342474

**Authors:** Jessica K Bone, Feifei Bu, Daisy Fancourt, Daniel Hayes

## Abstract

**Background:** Preliminary evidence indicates that social prescribing (SP) can improve children and young people’s (CYP) wellbeing but is limited by small non-representative samples and often relies on descriptives statistics. Given the wide implementation of SP in the UK, administrative records provide a unique opportunity to understand current practice and assess impacts on wellbeing.

**Objectives:**

1. To understand the quality of data captured in SP administrative records
2. To explore which CYP are currently receiving SP and what SP entails in practice
3. To assess the impact of SP on wellbeing

**Methods:** We used administrative records from one CYP SP service in England. Records were extracted from Joy, an online platform for managing SP. Over 18 months, 770 age-eligible CYP were referred to SP, 203 of whom were successfully discharged and completed two measures of wellbeing (the short Warwick-Edinburgh Mental Wellbeing Scale; SWEMWBS) at least seven days apart. We used descriptive statistics, a paired t-test to assess changes in wellbeing, and linear regressions with interactions to test effect modification.

**Findings:** Missing data was the largest issue, with ethnicity missing for 94% of CYP. A lack of detail and inconsistent recording also presented challenges. Despite this, we identified that most CYP were referred by their GP, followed by their school, with 97% referred because of their mental health. The most common pathway was to receive SP for less than 100 days, with 10-15 link worker contacts, and six contact hours. Following SP, SWEMWBS scores improved by 3.72 points (t(202)=17.50, 95% CI=3.30 to 4.14, p<0.001), a 20% relative increase. Exploratory analyses suggested that this increase was greater for those with fewer link worker contacts, no interventions recorded, and lower baseline wellbeing.

**Conclusions:** Despite numerous challenges with missing data and data quality, we found that CYP wellbeing increases following SP (as it is currently implemented). Effect sizes were consistent with larger studies of adults.

**Clinical implications:** Further development of online platforms is needed to monitor access to, nature of, and efficacy of SP. For those working in SP, we recommend more training, implementation of standardised guidelines, and designated time to update records.

**Key messages:** *What is already known on this topic:*

- Social prescribing has been widely implemented in the UK, but there is very little evidence on what it looks like or whether it works for children and young people

*What this study adds:*

- We show that it is feasible to evaluate social prescribing for children and young people using administrative records and identify several priorities for improving data quality
- We describe the typical social prescribing pathway in one children and young people’s service in England, which includes receiving SP for less than 100 days, with 10-15 link worker contacts, and 6 contact hours
- We found evidence for clinically significant improvements in children and young people’s wellbeing from their first to their last social prescribing session

*How this study might affect research, practice or policy:*

- To meet best practice guidelines for recording and evaluating social prescribing, we need further collaboration between online platform providers and social prescribing services, more training and time for link workers, and for research and policy to emphasize the importance of measuring outcomes

## Background

Social prescribing (SP) is a care pathway that aims to connect people with non-medical forms of support within their community, based on their values and preferences.^1^ It aims to address people’s social, emotional, and practical needs, which are often closely related to their medical needs but not routinely addressed by clinical treatments.^2^ In the UK, SP typically consists of referral to a link worker, or other similar professional, who works with the individual to develop a personalised care plan that connects them to community support. This may include a wide range of activities such as sports, arts, volunteering, counselling, housing support, training, and employment advice.^5^

SP could reduce the impact of social inequalities on children and young people (CYP), with potential for sustained benefits throughout the life course.^6,7^ Yet, despite being funded as an all-age approach in the UK,^8^ SP research, policy, and practice have predominantly focused on adults.^6,9,10^ This is particularly problematic because SP approaches for adults may not be suitable for CYP, who have different social contexts^11^ and face different challenges, including 75% of mental health problems emerging during adolescence.^12^ Additionally, CYP and adults may access SP through differing routes in the UK. Unlike adults who are primarily referred by GPs, CYP may access SP via alternative referral routes, including educational institutions and social care.^9^

Preliminary evidence among CYP is promising, with six uncontrolled pre-post quantitative studies^13–18^ and one randomised controlled trial (RCT)^19^ reporting improved wellbeing, reduced loneliness, and less healthcare utilisation following SP. But only one of these included over 100 participants in outcome analyses^19^ (n=6 to n=77 in the others),^13–18^ meaning most were likely underpowered. A review concluded that research with CYP has been further limited by not being representative of the target population, no control groups, short follow-ups, attrition, limited use of standardised measures and statistical analyses, and unclear reporting.^6^

Large RCTs of SP for CYP are clearly needed, and several are underway.^20,21^ But data capturing the widespread implementation of SP are also important.^9,22^ Administrative data are uniquely placed to evaluate real-world heterogeneity, allowing us to explore how CYP are currently accessing SP, whether there are inequalities in who receives SP, and whether SP (as currently implemented) can improve wellbeing. Link workers in the UK often use electronic health systems like EMIS or SystmOne, but many use bespoke systems designed for SP,^23^ including Joy,^24^ Elemental,^25^ and Social Rx Connect.^26^ These platforms allow link workers to monitor referrals, connect with services, record appointments and prescriptions, and measure impact. These platforms only include individuals who receive SP, meaning no control group is available for comparative analyses. However, they capture non-primary care referrals, provide rich data on interventions, and measure outcomes, providing major advantages over GP records. A growing body of research using administrative records to evaluate SP has emerged,^5,27–31^ but no studies have focussed on CYP.

## Objectives

We had three objectives. First, to understand the quality of administrative data captured in a platform for SP with CYP. Second, to explore which CYP are receiving SP, and what SP looks like for them. Finally, to assess the impact of SP on CYP wellbeing. Findings were used to make recommendations on how electronic health records should capture SP and identify targets for improving practice.

## Methods

### Data

We used administrative data from one SP service for CYP aged 11-18 years (or up to 25 with special educational needs and disabilities) in one county in North East England. This county includes diverse areas, with 5-10% of the population living in areas in the most deprived Index of Multiple Deprivation (IMD) decile and approximately 20% in the least deprived decile.^32^ The service used Joy,^24^ an online platform for healthcare professionals to manage SP, monitor clients, connect with local services, and measure impact. Joy is used in 30% of primary care networks, with 25,000 patients referred through Joy each month, and >19 million patients to date.^24^ Administrators and link workers used Joy to record referral sources and reasons, demographic information, contacts with individuals, prescribed activities and services, outcomes, discharge, and case notes. Information is entered through a combination of dropdown lists and free text boxes.

Data were available from Joy for March 2023 to September 2024. In this period, a total of 780 people were referred to SP (Figure 1). Of these, 770 were age-eligible for SP, and formed our analytical sample, in which we explored the characteristics of young people referred to SP.

**Figure 1.**
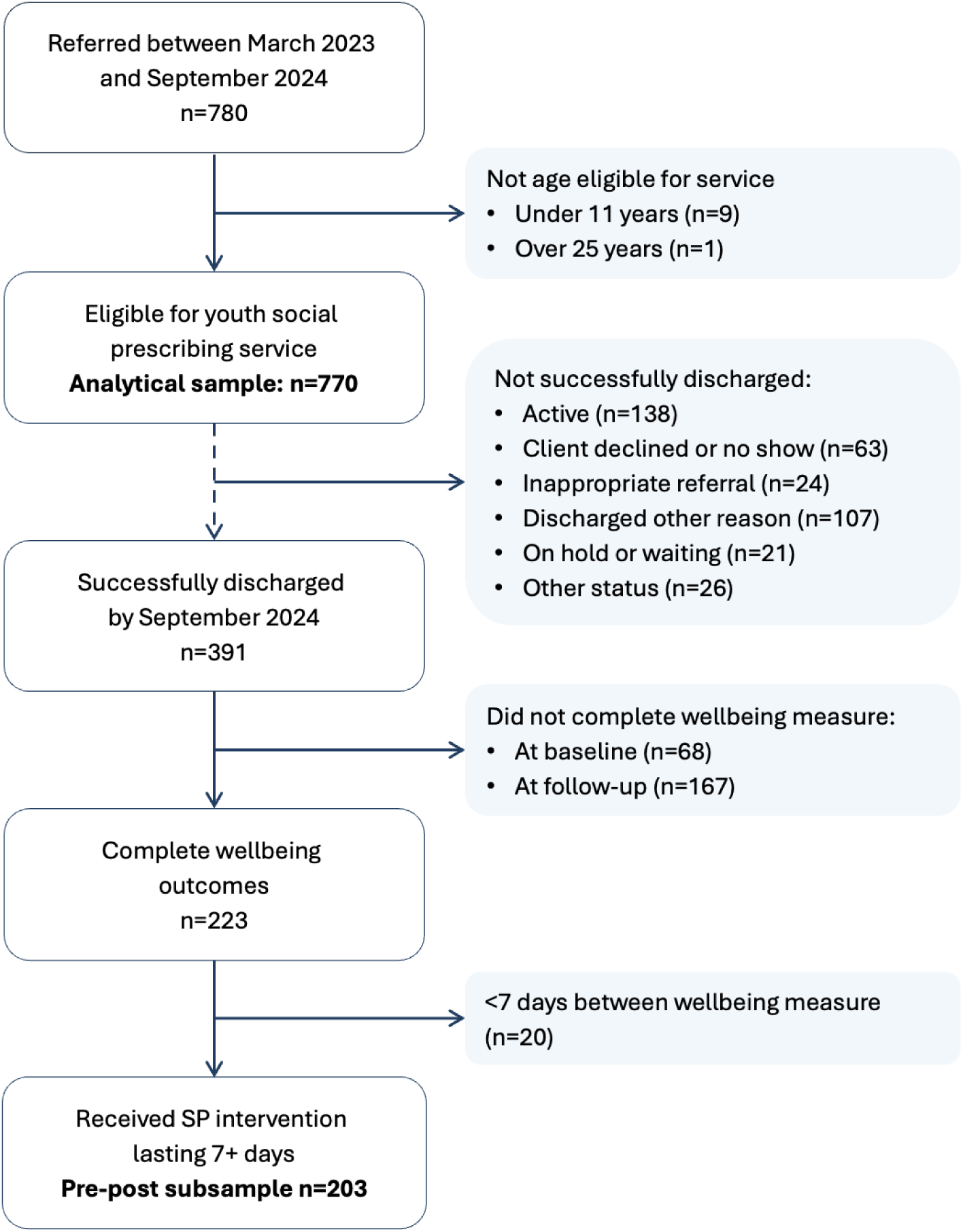
Flowchart showing sample selection.

We conducted further analyses among a subsample with data on wellbeing before and after receiving SP. To be included, individuals had to be successfully discharged from the SP service by September 2024 and have completed two measures of wellbeing. These outcome measures were typically completed at the first SP session and the final session before discharge. We also required that the final measure of wellbeing was completed at least seven days after the first wellbeing measure, ensuring that all individuals received at least one SP session and maximising sample size. Of the 770 eligible for SP, 391 were successfully discharged, 223 of whom had complete wellbeing outcomes, and 203 received a SP intervention lasting seven or more days. Our pre-post sample thus included 203 people, which had over 90% power to detect a mean difference of one standard deviation in wellbeing (based on UK population norms)^33^ using a paired t-test with alpha=0.05. The average length of time between wellbeing measures was 89.7 days (SD=56.9).

Analysis of this routine data was approved by UCL Research Ethics Committee (12467/006).

### Measures

#### Individual characteristics

Administrators recorded clients’ age, gender, and ethnicity based on their referral forms. We could not use gender and ethnicity due to the large proportion of missing data (92-94% missing; Table S1).

Young people were referred to SP from a range of sources, recorded using free text, which we collapsed into five categories: GP, school, other medical services (e.g. A&E, public health nursing services), youth services, or self-referral. Reasons for referral were recorded using >140 options, which we categorised into eight domains: mental health, physical health and wellbeing, social relationships, family issues, lifestyle, education employment and training, practical support, and other reasons (e.g. abuse, gender identity; Table S2). Following previous analyses of SP records,^5^ grouping was done independently by two authors with disagreements (10%) discussed and resolved by consensus. Multiple referral reasons were allowed.

The short Warwick-Edinburgh Mental Wellbeing Scale (SWEMWBS) measured mental wellbeing, with raw scores transformed to metric scores on the interval scale. Higher scores indicated greater wellbeing (range 7-35).^34^ The SWEMWBS has been validated for ages 15-21,^35,36^ and the longer WEMWBS for 13 and above.^37^ As some individuals did not understand specific words in the SWEMWBS (e.g. optimistic) or the rating scale, the SP service adapted it by adding emojis representing response options. It was completed with the link worker, who asked and explained the questions and recorded responses. The SWEMWBS was typically done in CYP’s first session (baseline), although sometimes there was a delay in completing it due to the waiting list, room availability, or the CYP’s willingness to engage. The same process was then repeated in CYP’s final session before discharge (follow-up).

#### Social prescribing

Link workers recorded the date of their first contact with the client, which could be an introductory appointment, text, or phone call. A first contact form was then completed at the first link worker appointment. However, as this form was only complete for 36% (n=140) of individuals who were successfully discharged, we could not use it as an indicator for engaging with SP. We therefore calculated the length of the SP period as the number of days from the first recorded contact to the final SWEMWBS completion (which should be done during the final session before discharge).

We were interested in the number of contacts each individual had with a link worker and the length of time each link worker spent with the client overall (number of contact hours). Contacts could include face-to-face or online meetings, texts, or emails; this information was not available. Contacts may lead to individuals being connected with community resources, either through onward referrals or through signposting, which we hereby refer to as receiving an intervention. We used the number of interventions recorded by the link worker. However, a lack of recorded interventions does not necessarily mean that young people did not receive any onward referrals or signposting.

Over 70 intervention types were reported. We classified them into five domains: mental health and wellbeing support, community activities, practical support, special educational needs and disabilities (SEND) support, or other services (Table S3). Domains were not mutually exclusive, as people were prescribed multiple interventions. Intervention domain was missing for 33% of the recorded interventions.

### Statistical analysis

We first summarised age, referral reasons, and referral sources for the full analytical and pre-post samples. Next, we described the SP pathway for the pre-post sample, including number of link worker contacts, contact hours, and types of interventions prescribed. We then evaluated SP using a two-tailed paired t-test, assessing whether there was a reduction in SWEMWBS scores from baseline to follow-up. Assumptions of the paired t-test were met, with differences of the paired values normally distributed.

In exploratory supplementary analyses, we tested whether changes in wellbeing were influenced by individual characteristics and the SP pathway. We dichotomised all potential effect modifiers to maximise group sizes and aid interpretation, using a median split for age (12-15 vs 16-20 years), number of contacts (1-13 vs 14-72), and number of contact hours (0.8-6.0 vs 6.1-33.0). We also compared those with no interventions recoded to those with one or more interventions recorded. For wellbeing at baseline, we used a SWEMWBS score of 18 or less to indicate probable clinical depression.^33,38^ This threshold has been developed in adults through comparison to the Patient Health Questionnaire (PHQ-9), on which a sore of 10 or more indicates clinical depression with a sensitivity of 88% and a specificity of 88%.^39^ We then used linear regression models testing the association between timepoint and SWEMWBS score, with interactions between timepoint and each of the potential effect modifiers in separate models. Cluster-robust standard errors accounted for repeated measures within individuals.

## Findings

### Eligible individuals

Among the 770 eligible clients referred to SP, ages ranged from 11 to 25 years (mean=15.4, standard deviation [SD]=2.3; Table S4). Most individuals were referred by their GP (69%) or school (22%), with relatively few referred by other medical services (5%), youth services (3%), or self-referral (1%).

Overall, 42% of individuals were referred for multiple reasons. Looking at referral reasons separately, mental health was the most common (listed for 97% of people), followed by social relationships (13%), and other reasons (13%; Figure 2A). Family issues was the least common domain (4%). Referral rate for practical reasons was similar (4%), but this was never used as the sole referral reason. Of those referred for mental health reasons, 41% also had other referral reasons.

**Figure 2.**
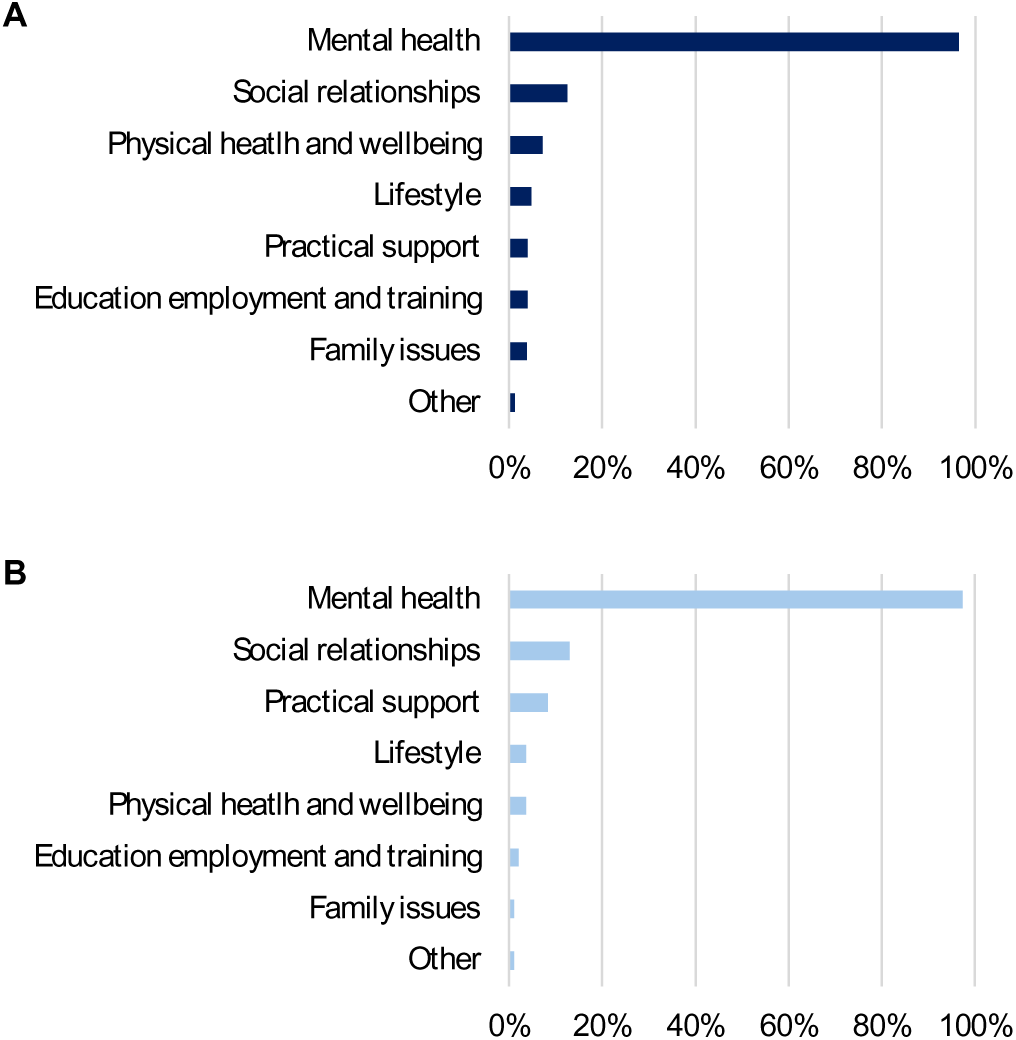
Referral reasons for the analytical sample (n=740; n=30 missing) and the pre-post sample (who were successfully discharged and completed outcome measures; n=192; n=11 missing). Domains were not mutually exclusive, allowing young people to have more than one referral reason.

Among all eligible referrals who completed the SWEMWBS at their first session (n=527), the mean score was 18.83 (SD=2.76). Among those referred for their mental health, 47% met the cut-off for probable clinical depression on the SWEMWBS. In contrast, 67% of those who did not have mental health listed as a referral reason met this threshold.

### Individuals who completed social prescribing

The 203 CYP in our pre-post sample were aged 12 to 20 (mean=15.2, SD=2.0), with 74% referred by their GP, 22% by school, and <5% by other routes. None were self-referrals (Table S4). Overall, 34% were referred for multiple reasons, with mental health the most common reason (97%), followed by social relationships (13%), and other reasons (8%; Figure 2B). Practical support and family issues were least common (<5%). In this subsample, the mean baseline SWEMWBS score was 18.74 (SD=2.55).

#### Social prescribing pathways

In this pre-post sample, link workers recorded between 1 and 72 contacts with clients (mean=16.7, SD=11.8, median=13; Figure 3A), over a period of 7-490 days (mean=104.7, SD=61.7, median=93.0). Contact hours ranged from 0.8 to 32.7 hours, with a mean of 6.7 hours (SD=4.0, median=6.0). Although there were outliers, the most common SP pathway was to receive SP for less than 100 days, with 10-15 link worker contacts (which could include texts and emails as well as meetings), and six contact hours (Figure 3B).

**Figure 3.**
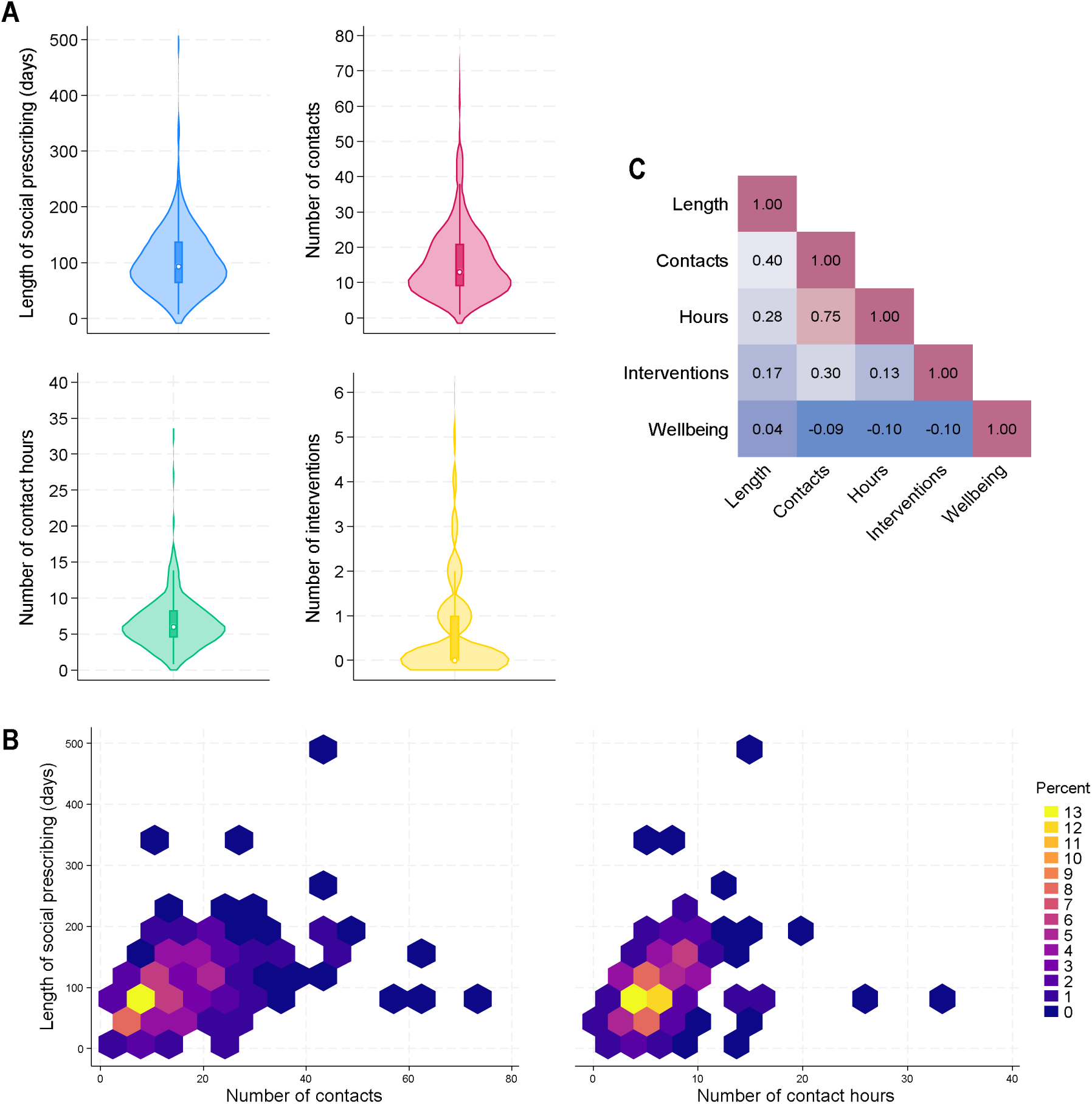
Descriptive statistics for social prescribing pathways in the pre-post sample (n=203). A) Violin plots showing distribution of the length of social prescribing, number of contacts with a link worker, number of contact hours, and number of recorded interventions. B) Hexagon heat plots showing the distribution of number of contacts and number of contact hours by the length of the social prescribing period. C) Heat plot of the correlations between social prescribing characteristics and baseline wellbeing (SWEMWBS score).

The length of the SP period was moderately positively correlated with the number of contacts (r=0.40; Figure 3C), but less strongly related to the number of contact hours (r=0.28). The number of contacts and contact hours were strongly correlated (r=0.75). But these SP characteristics were only weakly related to young people’s baseline wellbeing (r=-0.10 to 0.04). Looking at referral reasons, those with a single referral reason had more contacts (mean=17.6, SD=12.0) and more contact hours (mean=7.1, SD=3.7) than those with multiple referral reasons (contacts=14.2, SD=11.3; hours=5.8, SD=4.4).

No intervention was recorded for 63% of CYP (Figure 4A). For those with interventions recorded (n=76), number of interventions ranged from 1 to 6 (mean=1.9, SD=1.2). Intervention domain was reported for 68% of all recorded interventions, with mental health and wellbeing support most common (66%), followed by practical support (24%), and community activities (21%). SEND support (16%) and other services (3%) were least common.

**Figure 4.**
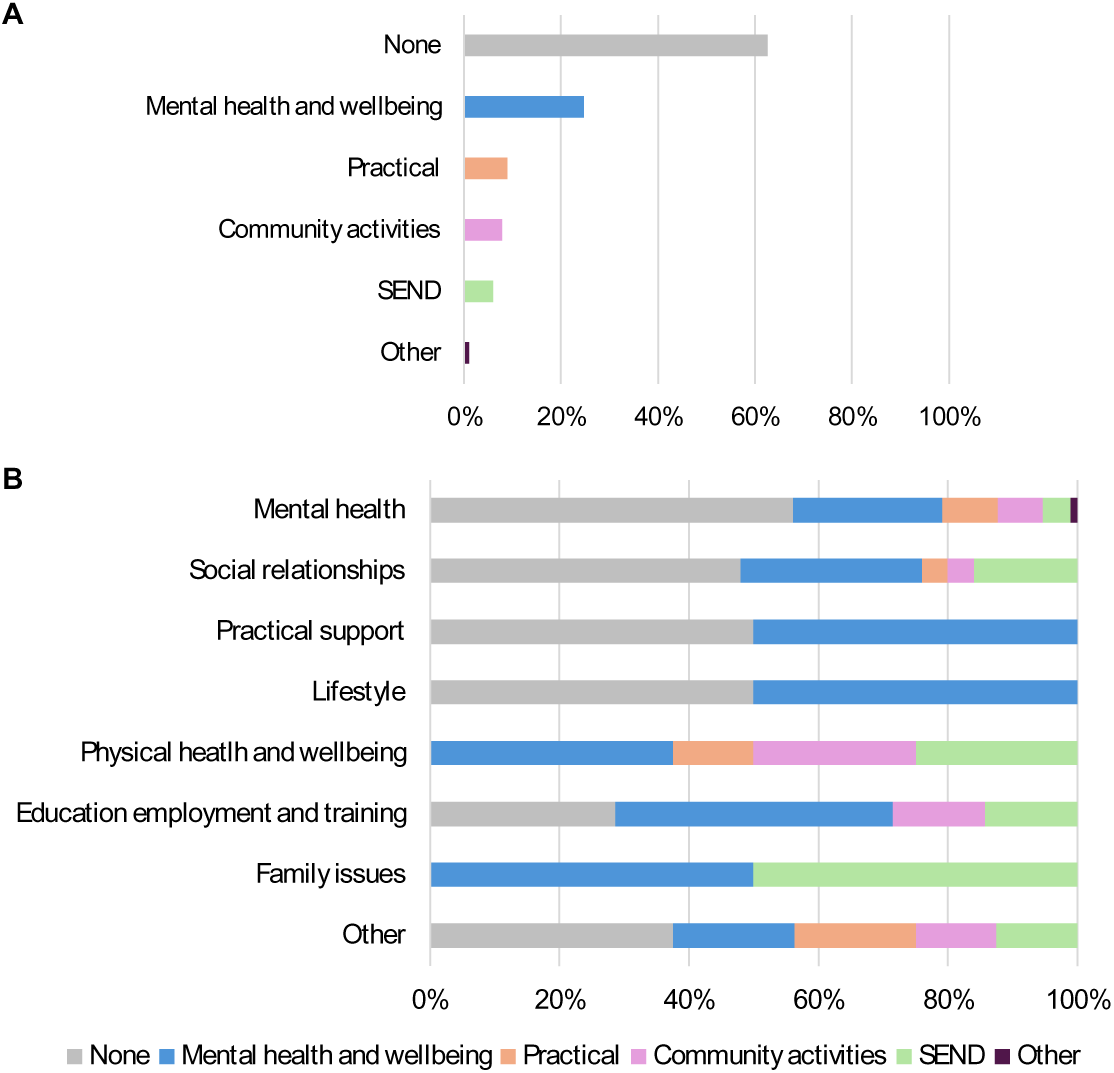
Intervention domains for A) those who were successfully discharged and completed outcome measures (the pre-post sample; n=203) and B) for the same group, shown separately according to their reasons for referral (group size differed across referral reasons). Domains were not mutually exclusive, as young people may have been referred/signposted to more than one intervention.

We then explored whether prescribed interventions corresponded to referral reasons (Figure 4B). Of those referred because of mental health, 56% had no intervention recorded. Similar proportions of those referred for support with lifestyle (50%), practical issues (50%), and social relationships (48%) had no intervention recorded. In contrast, all of those referred for family issues and physical health had an intervention recorded, most frequently a mental health and wellbeing intervention (family issues=50%, physical health=38%). However, these proportions were likely influenced by small and differing group sizes across referral reasons (n=2 to 187).

#### Efficacy of social prescribing

A paired t-test indicated that, following SP, SWEMWBS scores improved by an average of 3.72 points (t(202)=17.50, 95% CI=3.30 to 4.14, p<0.001, d=1.23). This is equivalent to a 20% relative increase in wellbeing from the first to final session. At the final session before discharge, the mean SWEMWBS score was 22.46 (SD=3.28).

In exploratory analyses, there was evidence for effect modification by the number of contacts (Figure 5B) and whether young people were prescribed interventions (Figure 5D). Young people with 14-72 contacts had a smaller increase in wellbeing than those with 1-13 contacts (linear regression interaction coef=-1.21, 95% CI=-2.04, -0.40, p=0.004). Young people with one or more prescribed interventions recorded also had a smaller increase in wellbeing than those with no interventions recorded (coef=-1.01, 95% CI=-1.87, -0.15, p=0.021). There was also evidence that the effect of SP differed according to wellbeing at baseline (Figure 5E), with a larger increase in wellbeing among those who met the criteria for probable depression at baseline compared to those who did not (coef=1.31, 95% CI=0.49, 2.13, p=0.002). However, across all subgroups, SWEMWBS scores still improved over time. There was no evidence for effect modification by age or contact hours (Figures 5A, 5C; Table S5).

**Figure 5.**
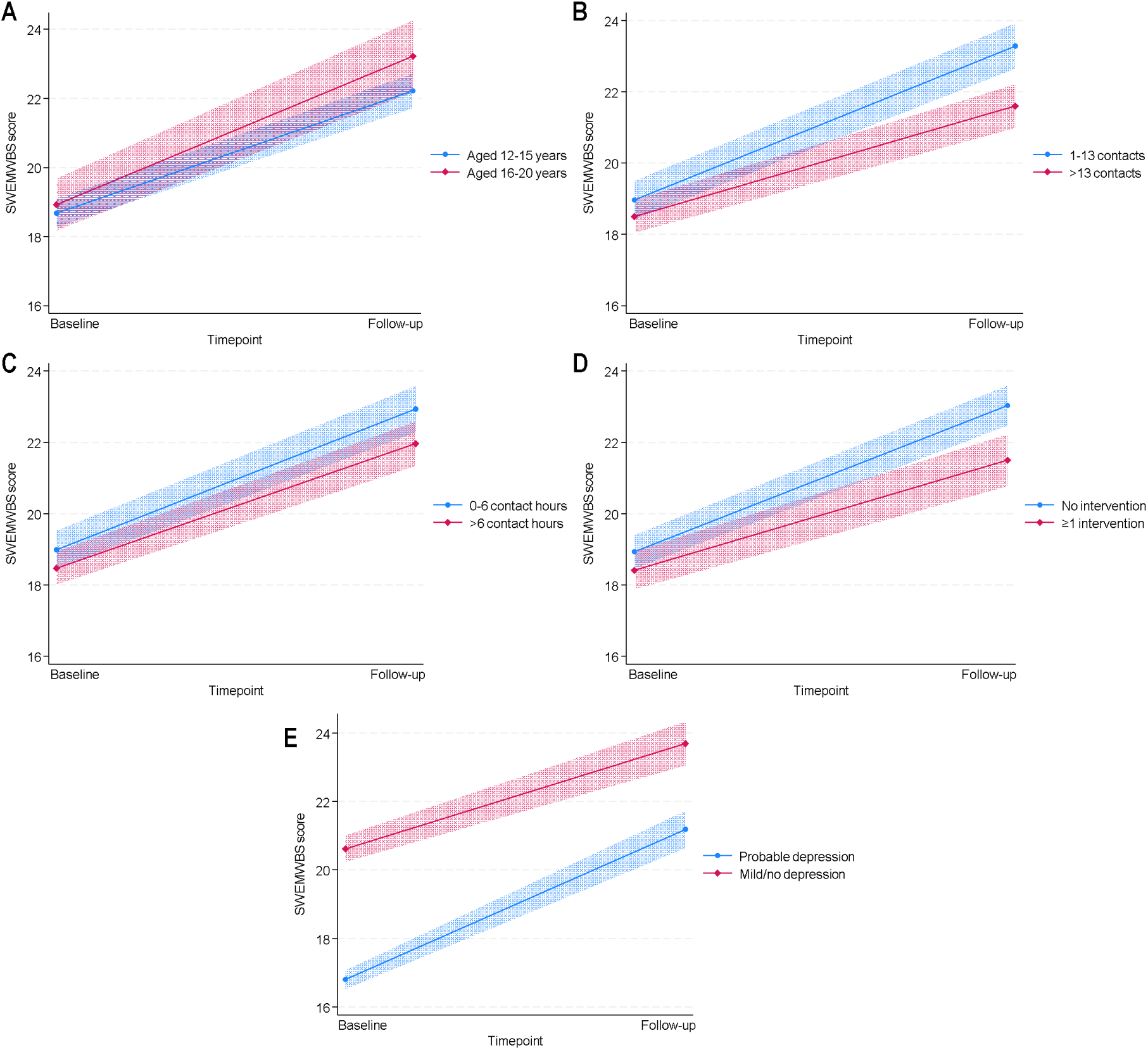
Exploratory analyses testing whether the change in wellbeing over time differed according to individual and social prescribing characteristics in the pre-post sample (n=203). Estimates from linear regression models with an interaction between timepoint and A) age group, B) number of contacts, C) contact hours, D) recorded interventions, and E) whether individuals met the criteria for probable clinical depression at baseline. Confidence intervals use cluster-robust standard errors, which account for the repeated measures within individuals.

## Discussion

We explored the feasibility of using routine data from the Joy platform^24^ to evaluate a SP service in North East England. Over 18-months from 2023 to 2024, 770 eligible 11 to 25-year-olds were referred and 391 were successfully discharged. Only 203 could be included in analyses of the impact of SP on wellbeing, largely due to missing data on wellbeing. Data quality precluded detailed analyses of inequalities in access to SP and the care pathway. However, we saw an increase in wellbeing from the first to the final session of SP. Exploratory analyses suggested that this increase differed according to the number of link worker contacts, number of interventions recorded, and initial levels of wellbeing.

Our first aim was to understand data quality. Missing data was the largest issue. Only 6-8% of CYP had gender/ethnicity recorded, so these could not be used. Just 57% of those successfully discharged had complete wellbeing measures, increasing the risk of selection bias. Only 36% of CYP who received SP had an intervention recorded, but no recorded interventions did not mean that no intervention was prescribed, so analyses using the number of interventions may not be valid. A lack of detail in some fields also presented challenges. We could not determine why some CYP were inappropriate referrals or discharged early. Understanding how many SP sessions each person received was difficult as we could not distinguish between contacts to arrange appointments and sessions. Additionally, link workers could not record re-referrals, meaning some cases appeared to be open for a long period, with very high numbers of sessions and contact hours, which may have resulted from multiple referrals.

Data limitations create issues for policy, practice, and research. The NHS England SP Information Standard recommends that patient demographics, needs and concerns, support offered (including referrals), and outcomes are recorded using the Office for National Statistics wellbeing measures (ONS4) or WEMWBS.^40^ The National Academy for SP (NASP) similarly recommends that interventions are recorded and a validated health/wellbeing questionnaire used to allow process and impact evaluation.^41^ In conversations with the service that provided these data, it emerged that gender and ethnicity were not compulsory fields, which likely contributed to high levels of missingness. In a 2025 survey of UK link workers, 24% said they never recorded outcome measures.^23^ Barriers to recording outcomes included a lack of appropriate sensitive and standard measures, little guidance on who should record outcomes or when and how to use measures, fragmented systems, and time constraints.^23^

To improve practice, we recommend further collaboration between platform providers and SP services. Services should be able to mandate fields, record why individuals are not eligible for SP or are discharged, identify re-referrals, and distinguish between contact types. NHS England recommends that SP systems link information from the national database of NHS patient demographics. Platforms currently differ in the functions offered, so SP services may wish to consider whether these are possible before choosing a platform. For those delivering SP, better digital tools, more training, standardised guidelines, reminders, and designated time for updating client records are needed.^23^ Further discussions may be needed around the utility of outcome measurement, particularly as 74% of link workers in the survey did not know whether outcomes were used to influence investment decisions.^23^

Our second aim was to explore access to and characteristics of SP for CYP. This was challenging due to the data limitations. Most CYP were referred by their GP or school. Referral reasons were very different to previous findings in adults, a third of whom were referred for mental health (vs 97% of CYP) and a quarter for practical support (vs 4% of CYP).^5^ This could be because this CYP SP service was originally commissioned to support mental health, although it does not specialise in mental health now. In our pre-post sample, the most common pathway was to receive SP for less than 100 days, with 10-15 link worker contacts, and 6 contact hours. However, there was wide variation in the SP period (up to 490 days), contacts (up to 72), and hours (up to 33). Link workers reported that some of extreme values may be due to safeguarding concerns or complex needs, where SP likely was not appropriate but they were not comfortable discharging people. However, most were likely driven by re-referrals. For the minority with intervention domains recorded, the most common were mental health and wellbeing support (66%), practical support (24%), and community activities (21%). Prescribed interventions did not match referral reasons well. Although it is hard to draw conclusions given the proportion of missing data, some of this disparity may be due to the explicit focus on what matters to individuals in SP. This may mean that interventions are well aligned CYP’s own perceived needs, which are not necessarily why they were referred. Baseline wellbeing, which was worse among those not referred due to their mental health, also indicates that referral reasons may not accurately represent CYP’s current state.

Our final aim was to assess the impact of SP on wellbeing. We found an average relative increase in wellbeing of 20% from the first to the last SP session. This corresponded to statistics provided to the SP service in a Joy monitoring dashboard. Whilst clinically meaningful,^42^ this is <4 SWEMWBS points, which sounds less significant than a 20% increase. It is thus important to present absolute alongside relative outcomes, and for dashboards to describe how statistics are calculated, including the underlying number of clients. Until then, practitioners cannot rely on dashboards to make informed decisions about services. Despite limitations, our findings are consistent with other evaluations. Uncontrolled studies among CYP have found similar pre-post increases in wellbeing in analyses of the SWEMWBS,^13^ ONS4,^13,18^ and descriptive statistics.^14,15,17^ National studies including all ages have found similar evidence, with analyses of records from Elemental showing a 3.31-point increase in SWEMWBS score^27^ and an evaluation of green SP (focussed on nature-based activities) finding small but robust increases on the ONS4.^43^

Our findings indicate some ways to optimise SP. In exploratory analyses, CYP with more link worker contacts (>13) or with 1+ interventions recorded had smaller increases in wellbeing. Although contacts include calls and messages alongside SP sessions, this could reflect case complexity or limited impact of SP beyond the recommended 6-12 sessions.^44,45^ SP is a short-term pathway. Where individuals receive many sessions, the care mechanism may not be working as intended, potentially because SP is not currently appropriate. This suggests that, if CYP have not responded within 12 sessions, practitioners should consider discharging them (if safe to do so), to avoid preventing others from accessing the service. In contrast, we found that those with lower wellbeing before starting SP experienced larger increases in wellbeing following SP. This is consistent with other evidence,^13,14^ and could be because these CYP had greatest room for improvement or because SP has greater impact for those with the worst mental health (who are still eligible and able to engage).

This study had several strengths, including being the first to use routine data to evaluate CYP SP. This enabled measurement of real-world heterogeneity and identification of strategies to improve data collection and quality. Wellbeing was measured with the SWEMWBS, which has been validated for this population^37^ and used in many evaluations of SP.^6,27,46^ However, 43% of CYP successfully discharged from SP had missing wellbeing data. Although the pre-post subsample was comparable to those referred to SP in terms of average age, referral routes/reasons, and wellbeing, this may have introduced selection bias in unmeasured characteristics. Additionally, missing outcome data limited the size of our pre-post sample, which reduced power for exploratory analyses of effect modification. We could not use multiple imputation because of the lack of demographic information.

Another major limitation is the lack of control group, as all CYP received SP. Our findings could thus reflect regression to the mean, meaning low wellbeing scores may appear to improve without any intervention.^47^ Given that no control group can be derived from SP records, future research should consider using synthetic controls^48^ or digital twins.^49^ However, detailed individual-level demographic and socioeconomic information is needed for these approaches. It is thus not yet feasible to create matched controls. This lack of information also precludes further analyses of inequalities in access to and engagement with SP. Finally, data quality meant that analyses of the number of interventions may reflect link workers’ recording practices rather than actual interventions received.

## Clinical implications

We have identified the challenges of using routine data to describe and evaluate SP for CYP. Despite numerous data limitations, we found that SP, as currently implemented in one area, is associated with improvements in CYP wellbeing. Our analyses suggest that SP may work best as a short-term care pathway, as recommended in best practice guidelines. We identified key priorities for enhancing data quality. Bespoke electronic records providers should consider compulsory recording of demographics, the ability to identify re-referrals, and clear recording of SP sessions. To support those working in SP, we recommend more training, better tools for recording outcomes, standardised guidelines and processes, and designated time for updating records. Finally, it is vital that policymakers and researchers continue to advocate for high quality data, particularly outcome measures.

## Data Availability

The individuals included in this evaluation did not give written consent for their data to be shared publicly, so due to the sensitive nature of the research supporting data are not available. Code for the analyses is available from JKB on request. We thank the Social Prescribing Service for granting us data access and for the numerous conversations and input that enabled this evaluation.

## Acknowledgements

We thank the Social Prescribing Service for granting us data access and for the numerous conversations and input that enabled this evaluation. This work was supported by the Economic and Social Research Council (ESRC) [UKRI1717] and the National Academy for Social Prescribing (NASP). JKB is also funded by a National Institute for Health and Care Research (NIHR) Advanced Fellowship (NIHR305289). This publication is independent research that was also supported by the National Institute for Health and Care Research ARC North Thames. The views expressed are those of the authors and not necessarily those of the NIHR or the Department of Health and Social Care.

## Data availability

The individuals included in this evaluation did not give written consent for their data to be shared publicly, so due to the sensitive nature of the research supporting data are not available. Code for the analyses is available from JKB on request.

## Supplementary Materials

**Table S1.** Missing data on study variables.

| Measure | Analytical sample<br>(n=770) | Pre-post<br>sample (n=203) |
| --- | --- | --- |
|  | N (%) missing |  |
| Age | 0 | 0 |
| Gender | 711 (92%) | 187 (92%) |
| Ethnicity | 721 (94%) | 190 (94%) |
| Referral source | 1 (<1%) | 0 |
| Referral reason | 30 (4%) | 11 (5%) |
| SP length | 548 (71%) | 0 |
| Number of contacts | 0 | 0 |
| Number of contact hours | 0 | 0 |
| Number of interventions | 0 | 0 |
| Intervention domain<br>(proportion of prescribed interventions) | 112 (33%) | 46 (32%) |
| Wellbeing (SWEMWBS) |  |  |
| Baseline | 243 (32%) | 0 |
| Follow-up | 511 (66%) | 0 |

*Note.* Dashes indicate no missing data.

**Table S2.** Examples of reasons for referral to social prescribing, which were recorded using free text.

| **Domain** | **Examples** |
| --- | --- |
| Mental health | anger and frustration; anxiety; bereavement; coping with change; depression; emotional wellbeing; feeling low; feeling unsafe; lack of motivation; low mood; panic attacks; previous self-harm, self-esteem; social anxiety; stress; suicidal ideation; traumatic life events |
| Physical health and wellbeing | ADHD; autism; coeliac disease; cognitive disorder; endometriosis; health and wellbeing support; high body weight; managing a long-term condition; neurodiversity; physical health; sexual health; Tourette's |
| Social relationships | concerns around socialisation; friends; loneliness/isolation; relationships; sex and relationships; struggling to make friends |
| Family issues | family difficulties; family problems; issues with family; relationship with family |
| Lifestyle | finding things to enjoy; problems with sleep; relationship with food; sedentary lifestyle; substance misuse; weight management |
| Education employment and training | education; employment; not supported at school; school attendance; school life; work/education; work/training/education |
| Practical support | caring responsibilities; day-to-day helping hand; finances; housing problem; money/benefits |
| Other | bullying; connecting to services; domestic abuse in the home; gender/sexual identity; in foster care; parental split; problems at home; transitioning gender; victim of abuse |

**Table S3.** Examples of interventions that people were referred to, which were recorded using free text.

| **Domain** | **Examples** |
| --- | --- |
| Mental health and wellbeing support | Bereavement Counselling; Calm Harm app; CAMHS; Community paediatric diabetes service; Headspace; IAPT; OCD Action; Samaritans; YoungMinds |
| Community activities | boxing club; disability football group; drama classes; horse riding; library reading group; mums and friends group; Scouts; tennis club; youth action group |
| Practical support | Citizens Advice; domestic abuse support services; food bank; safeguarding children partnership; weight management service |
| Special educational needs and disabilities (SEND) support | Early Help support; parent carer forum; SENDIASS; sensory processing hub |
| Other | meditation; mindfulness; sleep; small locally focused charity |

**Table S4.** Individual characteristics and description of the social prescribing pathway.

|  | **Analytical sample (n=770)** | **Pre-post sample (n=203)** |
| --- | --- | --- |
| **Individual characteristics** |  |  |
| Age (years) | 15.38 (2.25) | 15.24 (1.98) |
| Referral source |  |  |
| GP | 69% | 74% |
| School | 22% | 22% |
| Other medical services | 5% | <5% |
| Youth services | 3% |  |
| Self-referral | 1% | - |
| Baseline wellbeing (SWEMWBS) | 18.83 (2.76) | 18.74 (2.55) |
| **Social prescribing pathway** |  |  |
| Social prescribing length (days) | - | 104.66 (61.72) |
| Number of contacts | - | 16.71 (11.83) |
| Number of contact hours | - | 6.67 (3.97) |
| Number of interventions | - | 0.70 (1.17) |

**Table S5.** Linear regression models testing whether association between time and wellbeing score is modified by individual and social prescribing pathway characteristics in the pre-post sample (n=203).

| **Effect modifier** | **Coefficient (95% CI)** | **p value** |
| --- | --- | --- |
| Baseline wellbeing<br>(ref: no probable depression) | 1.31 (0.49, 2.13) | 0.002 |
| Age<br>(ref: 12-15 years) | 0.75 (-0.30, 1.81) | 0.161 |
| Number of contacts<br>(ref: 1-13) | -1.22 (-2.04, -0.40) | 0.004 |
| Number of contact hours<br>(ref: 0-6) | -0.46 (-1.29, 0.38) | 0.286 |
| Interventions prescribed<br>(ref: no interventions) | -1.01 (-1.87, -0.15) | 0.021 |

*Note.* Coefficients shown are for the interaction between the effect modifier and time.

